# Clinical deep sequencing to diagnose pathogenic mosaic variants in malformations of cortical development and epilepsy

**DOI:** 10.64898/2026.09.01.26361943

**Authors:** Katelyn Stone, Gillian Prinzing, Abbe Lai, Lacey Smith, Beth R. Sheidley, Meagan M. Corliss, Kevin M. Bowling, Yang Cao, Kimberly Wiltrout, Scellig S.D. Stone, Hart Lidov, Edward Yang, Annapurna Poduri, Alissa M. D’Gama

## Abstract

**Background and Objectives:** Deep sequencing of brain tissue in the research setting has established that mosaic variants are a major cause of malformations of cortical development (MCDs) and epilepsy. However, genetic testing in the clinical setting primarily detects germline variants using clinically accessible samples. We aimed to determine the diagnostic yield and clinical utility of deep sequencing in the clinical setting to identify pathogenic mosaic variants for this population.

**Methods:** We performed a retrospective cohort analysis of individuals at Boston Children’s Hospital with MCDs with or without epilepsy who received clinical deep sequencing between September 2017 and February 2026. Demographic, clinical, and genetic testing data were abstracted from the medical record. For individuals without systemic features, we classified brain tissue as an affected tissue sample. For individuals with systemic features, we classified brain or relevant non-brain tissue as affected. The primary outcome was the diagnostic yield of clinical deep sequencing performed using affected vs unaffected tissue samples. The secondary outcome was the clinical utility of genetic diagnoses.

**Results:** Our cohort included 37 individuals (19/37 (51%) female, 18/37 (49%) male) with MCDs, of whom 35/37 (95%) had epilepsy (25 with brain tissue samples available from epilepsy surgery) and 8/37 (22%) had systemic features. Most (35/37 (95%)) had dysplasia phenotypes on MRI and 12/27 (44%) with pathology available had Focal Cortical Dysplasia Type I or II. The diagnostic yield was 53% (17/32; 16 mosaic and 1 germline variant) when clinical deep sequencing was performed using an affected tissue sample vs 0% (0/6) using an unaffected tissue sample (p=0.016). Of the diagnosed cases, 13/17 (76%) had testing performed on brain tissue (1 with systemic features) and 4/17 (24%) on non-brain tissue (3 buccal and 1 duodenal tissue, all with systemic features). All but one diagnosis involved the mTOR pathway. All diagnoses had clinical utility.

**Discussion:** Clinical deep sequencing, when performed using an affected tissue sample, has high diagnostic yield and clinical utility for individuals with MCDs, especially dysplasia phenotypes, and epilepsy. Our findings support implementation of clinical deep sequencing for this population, especially as the genetic diagnoses have implications for emerging precision therapies.

## INTRODUCTION

Malformations of cortical development (MCDs) encompass a range of neurodevelopmental disorders with malformations classified into three groups: abnormal neuronal and glial proliferation or apoptosis (Group I), abnormal neuronal migration (Group II), or abnormal post-migrational development (Group III).^1^ Collectively, MCDs are a common cause of epilepsy, especially early onset drug resistant epilepsy (DRE), and developmental delay. Prior studies have reported MCDs in up to 25% of children and 10% of adults with epilepsy, with higher percentages reported in individuals with non-acquired epilepsy, focal epilepsy, and DRE.^2–6^ Most individuals with MCDs develop epilepsy, and for eligible individuals with MCD-associated DRE, surgical treatment remains standard of care.^7–10^ Based on histopathology of resected brain tissue in a large cohort of individuals with DRE who underwent surgery, MCDs represented the third most common diagnosis overall and in adults and the most common diagnosis in children.^11^

While non-genetic causes contribute to some MCDs, there has been growing recognition of the role of genetic causes—both germline and mosaic variants—in the genomic era. A germline variant is present in all cells of an individual and thus detectable in any tissue. Over the past several decades, inherited and *de novo* germline variants in multiple genes have been associated with MCDs and epilepsy.^12^ A mosaic variant arises post-zygotically and is present in only a subset of cells of an individual. Depending on the time and place of the somatic mutation event, a mosaic variant may be detectable across multiple tissues (e.g., in brain and non-brain tissues if it arose relatively early in development before gastrulation) or only in specific tissues (e.g., only in brain tissue if it arose relatively late in development during neurogenesis). Mosaic variant detection requires deep sequencing, optimized bioinformatic pipelines, and ideally orthogonal validation to discriminate mosaic variants from sequencing artifacts. Over the past decade, deep sequencing of brain tissue in the research setting has established that *de novo* mosaic variants, particularly brain-limited mosaic variants, are an important cause of MCDs and epilepsy.^13^ Multiple studies have demonstrated that mosaic variants that activate the mammalian target of rapamycin (mTOR) pathway lead to a spectrum of cortical dysplasias, notably Focal Cortical Dysplasia (FCD) Type II and hemimegalencephaly (HME).^14, 15^ Mosaic variants in *SLC35A2* were subsequently identified in individuals with mild malformation of cortical development with oligodendroglial hyperplasia and epilepsy (MOGHE).^16, 17^ Most recently, mosaic variants that activate the Ras/Raf/MAPK pathway have been associated with mesial temporal lobe epilepsy (mTLE).^18^

Although mosaic variants represent a major cause of MCDs and epilepsy, genetic testing in the clinical setting for individuals with MCDs and epilepsy primarily detects germline variants using clinically accessible samples.^19^ Thus, a gap exists between mosaic variant detection in the research setting and translating these methods to the clinical setting for this population. Here, we aimed to determine the diagnostic yield and clinical utility of performing deep sequencing in the clinical setting to identify pathogenic mosaic variants in individuals with MCDs and epilepsy.

## METHODS

### Standard Protocol Approvals, Registrations, and Individual Consents

The Boston Children’s Hospital (BCH) Institutional Review Board (IRB) approved this retrospective chart review study with a waiver of informed consent. A subset of the individuals included in this study, including those whose imaging is shown in Figure 3, are enrolled in BCH IRB-approved research studies for which the individual or their parent provided written informed consent. These include the Rosamund Stone Zander and Hansjoerg Wyss Translational Neuroscience Center Repository Core for Neurological Disorders, the Children’s Rare Disease Collaborative Unexplained Epilepsies study, and the Gene-STEPS study.^20, 21^

### Data Abstraction

We identified individuals with MCDs with or without epilepsy who received care at BCH and had clinical deep sequencing performed between September 2017 and February 2026 with the assistance of the genetic counselors who usually coordinated this genetic testing (GP and AL). Demographic, clinical, and genetic testing data were abstracted from the electronic medical record (EMR) through July 2026 and stored in a BCH-hosted Research Electronic Data Capture database.^22^ Demographic variables abstracted included sex assigned at birth and individual- or parent-reported race and ethnicity. Clinical variables abstracted, if applicable, included age of seizure onset, notable neurological and non-neurological clinical features, brain MRI findings, date of neurosurgery, and neuropathology findings. For each genetic test sent at BCH (and at other hospitals as available), we abstracted the type of test, date of sample collection, sample type, date of report, and results. For each genetic test, we determined whether the results led to a molecular genetic diagnosis. We defined a molecular genetic diagnosis as pathogenic or likely pathogenic (P/LP) variants, classified based on standardized criteria, that explained the individual’s MCD and, if applicable, epilepsy and other clinical features.^23–25^ We also determined whether diagnostic results had clinical utility in the categories of treatment/precision therapy, referrals, workup (laboratory tests and imaging), prognosis, and/or recurrence risk counseling. For clinical deep sequencing tests, we evaluated whether the sample used for testing represented an affected vs an unaffected sample. For individuals with MCDs without systemic (non-CNS) features, we considered brain tissue to represent an affected sample. For individuals with MCDs with systemic features (e.g., vascular malformation, systemic overgrowth), we considered brain or involved non-brain tissue (based on the reported systemic features) to represent an affected sample.

### Clinical Deep Sequencing Tests

The clinical deep sequencing tests performed included tests designed for individuals with suspected non-oncologic disorders of mosaicism and tests designed for individuals with known or suspected tumors.

The clinical deep sequencing tests designed for individuals with suspected non-oncologic disorders of mosaicism were performed through the Clinical Genomics Laboratory at Washington University School of Medicine in St. Louis and were usually sent after individuals were referred to neurogenetics for genetic counseling and test consent.^26^ The initial clinical deep sequencing test available through this laboratory was a Somatic Overgrowth (SO) panel (gene set updated over time; currently 49 genes). The laboratory subsequently expanded to offering a range of clinical deep sequencing panels based on phenotypes, including a Cortical Malformations and Epilepsy (CME) panel (for which AL, AP, and AMD assisted with gene selection and validation; currently 39 genes), which we started using in 2023.^27^ The clinical deep sequencing panels can be performed on frozen or formalin fixed paraffin embedded (FFPE) tissue samples and clinically accessible samples like blood or buccal swabs. The panels are targeted hybrid capture-based next-generation sequencing designed to detect single nucleotide variants (SNVs) and insertions and deletions (indels). At a mean depth of at least 4000x across the captured region, this test has 89.84% sensitivity for variants at a variant allelic fraction (VAF) of 1.25% and 99.48% for variants at a VAF of at least 2.5%.

The clinical deep sequencing tests designed for individuals with known or suspected tumors were performed through local laboratories and were usually sent at the discretion of the neurosurgeon and pathologist. The initial test available, Oncopanel (currently >400 genes), was performed through the Brigham and Women’s Hospital Center for Advanced Molecular Diagnostics.^28, 29^ This test, which can be performed on frozen or FFPE “tumor” samples and can include a comparator germline blood sample, can detect SNVs and indels at VAFs down to 10% and lower depending on depth/quality as well as copy number and structural variants (CNVs/SVs). In 2025, our institution switched to the BRIGHTseq Beacon test (currently >300 genes), which is performed through the BCH Laboratory for Molecular Pediatric Pathology.^30, 31^ This test, which is performed on an FFPE “tumor” sample and comparator germline blood sample, has an analytical sensitivity of 96.45% for SNVs at a VAF of 10% and 93.36% for indels at a VAF of 20% at a depth of at least 125x.

### Statistical Analysis

We analyzed summary statistics for clinical, demographic, and genetic testing features. We analyzed associations of these features with diagnostic clinical deep sequencing using chi-squared tests with statistical significance set at p<0.05.

### Data Availability

Deidentified demographic, clinical, and genetic testing data are available in the main and supplementary tables. Genetic variants were deposited in public databases like ClinVar per the policies of the clinical laboratories that reported the variants.

## RESULTS

### Study Cohort

Our cohort included 37 individuals with MCDs who received clinical deep sequencing, of which 19 (51%) were female and 18 (49%) were male. Individual- or parent-reported race and ethnicity were most commonly White (23/37 (62%)) and Non-Hispanic (29/37 (78%)), respectively. Of the 35 individuals with epilepsy, 22 (63%) had seizure onset in the neonatal/infantile period. Systemic features were present for 8/37 individuals (22%), including 3 individuals with megalencephaly-capillary malformation (MCAP) or MCAP-like syndrome, 1 with Congenital Lipomatous Overgrowth, Vascular Malformations, Epidermal Nevis, Spinal/Skeletal Anomalies/Scoliosis (CLOVES) syndrome, 1 with clinical features suggestive of PIK3CA-related overgrowth spectrum (PROS), 2 with dermatologic and size differences between the left and right sides of the body, and 1 with a renal angiomyolipoma. Regarding other comorbidities, 25/37 individuals (68%) had a history of developmental delay, 3/37 (8%) developmental regression, 7/37 (19%) autism spectrum disorder, and 5/37 (14%) attention deficit hyperactivity disorder.

MRI findings were suggestive of FCD, HME, or other complex dysplasia in 34/37 individuals (92%). Of the remaining 3 individuals, 2 had bilateral polymicrogyria (PMG) and 1 had microcephaly with simplified gyral pattern (MSGP). Pathology findings were available for the 27 individuals who underwent surgical resection/biopsy for DRE. Based on pathology, 12 individuals had definitive findings of FCD I or FCD II, including 2 with FCD IA, 1 with FCD IC, 6 with FCD IIA, and 3 with FCD IIB, based on International League Against Epilepsy classification.^32^ An additional 14 individuals had findings suggestive of MCD or other abnormal features, including 4 with MCD or mild MCD not otherwise specified, 5 with oligodendroglial hyperplasia (of which 1 also had periventricular nodular heterotopia and 2 also had hippocampal sclerosis), 1 with hyaline protoplasmic astrocytopathy, 2 with dysmorphic neurons, 1 with a neuronal migration defect, and 1 with mild increased white matter cellularity. One individual was found to have a low grade neuroepithelial tumor on pathology that was initially suspected to be an FCD on MRI. Demographic and clinical characteristics of the cohort are summarized in **Table 1**.

**Table 1:**
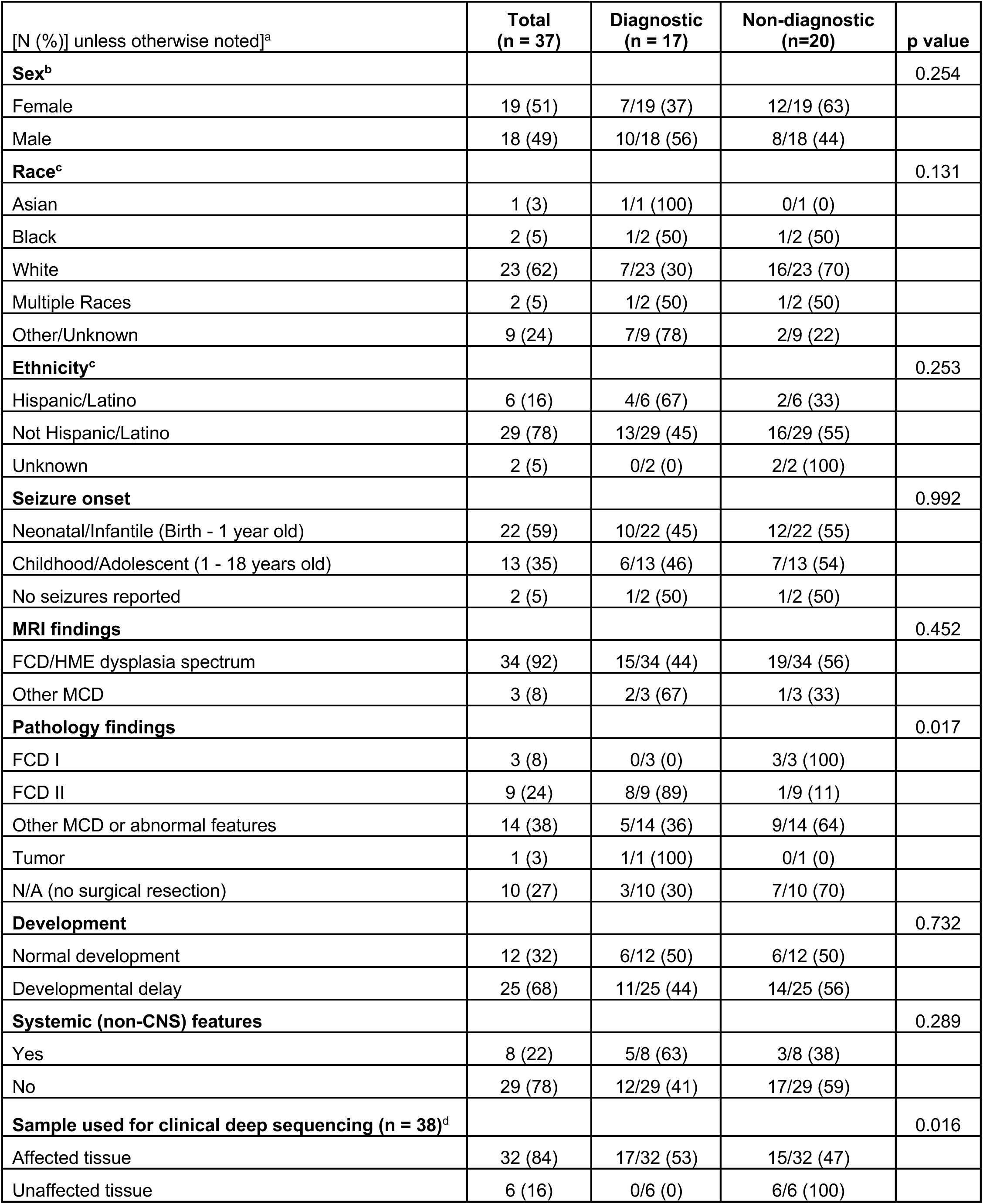
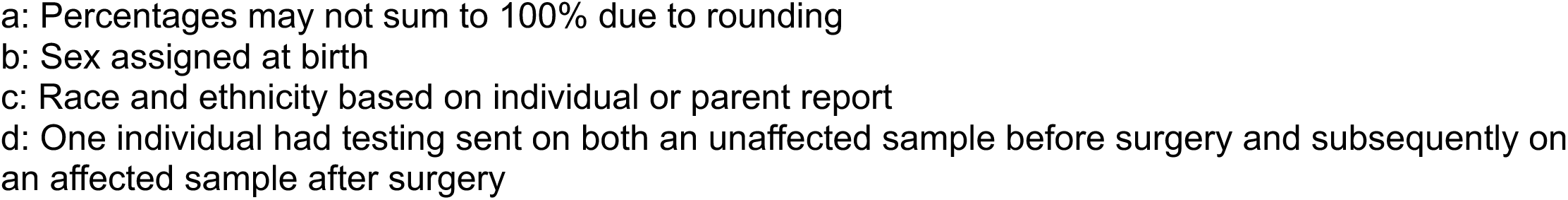
Summary of Cohort Demographic and Clinical Features.

| [N (%)] unless otherwise noted]<sup>a</sup> | Total<br>(n = 37) | Diagnostic<br>(n = 17) | Non-diagnostic<br>(n=20) | p value |
| --- | --- | --- | --- | --- |
| **Sex<sup>b</sup>** |  |  |  | 0.254 |
| Female | 19 (51) | 7/19 (37) | 12/19 (63) |  |
| Male | 18 (49) | 10/18 (56) | 8/18 (44) |  |
| **Race<sup>c</sup>** |  |  |  | 0.131 |
| Asian | 1 (3) | 1/1 (100) | 0/1 (0) |  |
| Black | 2 (5) | 1/2 (50) | 1/2 (50) |  |
| White | 23 (62) | 7/23 (30) | 16/23 (70) |  |
| Multiple Races | 2 (5) | 1/2 (50) | 1/2 (50) |  |
| Other/Unknown | 9 (24) | 7/9 (78) | 2/9 (22) |  |
| **Ethnicity<sup>c</sup>** |  |  |  | 0.253 |
| Hispanic/Latino | 6 (16) | 4/6 (67) | 2/6 (33) |  |
| Not Hispanic/Latino | 29 (78) | 13/29 (45) | 16/29 (55) |  |
| Unknown | 2 (5) | 0/2 (0) | 2/2 (100) |  |
| **Seizure onset** |  |  |  | 0.992 |
| Neonatal/Infantile (Birth - 1 year old) | 22 (59) | 10/22 (45) | 12/22 (55) |  |
| Childhood/Adolescent (1 - 18 years old) | 13 (35) | 6/13 (46) | 7/13 (54) |  |
| No seizures reported | 2 (5) | 1/2 (50) | 1/2 (50) |  |
| **MRI findings** |  |  |  | 0.452 |
| FCD/HME dysplasia spectrum | 34 (92) | 15/34 (44) | 19/34 (56) |  |
| Other MCD | 3 (8) | 2/3 (67) | 1/3 (33) |  |
| **Pathology findings** |  |  |  | 0.017 |
| FCD I | 3 (8) | 0/3 (0) | 3/3 (100) |  |
| FCD II | 9 (24) | 8/9 (89) | 1/9 (11) |  |
| Other MCD or abnormal features | 14 (38) | 5/14 (36) | 9/14 (64) |  |
| Tumor | 1 (3) | 1/1 (100) | 0/1 (0) |  |
| N/A (no surgical resection) | 10 (27) | 3/10 (30) | 7/10 (70) |  |
| **Development** |  |  |  | 0.732 |
| Normal development | 12 (32) | 6/12 (50) | 6/12 (50) |  |
| Developmental delay | 25 (68) | 11/25 (44) | 14/25 (56) |  |
| **Systemic (non-CNS) features** |  |  |  | 0.289 |
| Yes | 8 (22) | 5/8 (63) | 3/8 (38) |  |
| No | 29 (78) | 12/29 (41) | 17/29 (59) |  |
| **Sample used for clinical deep sequencing (n = 38)<sup>d</sup>** |  |  |  | 0.016 |
| Affected tissue | 32 (84) | 17/32 (53) | 15/32 (47) |  |
| Unaffected tissue | 6 (16) | 0/6 (0) | 6/6 (100) |  |

- a: Percentages may not sum to 100% due to rounding - b: Sex assigned at birth - c: Race and ethnicity based on individual or parent report - d: One individual had testing sent on both an unaffected sample before surgery and subsequently on an affected sample after surgery

### Genetic Testing and Diagnostic Yield

Clinical deep sequencing and genetic diagnosis for the cohort is summarized in **Figure 1**. Clinical deep sequencing was performed on an affected tissue sample for 32 individuals and on an unaffected tissue sample for 6 individuals (one individual had testing sent on both an unaffected sample before surgery and subsequently on an affected sample after surgery).

**Figure 1:**
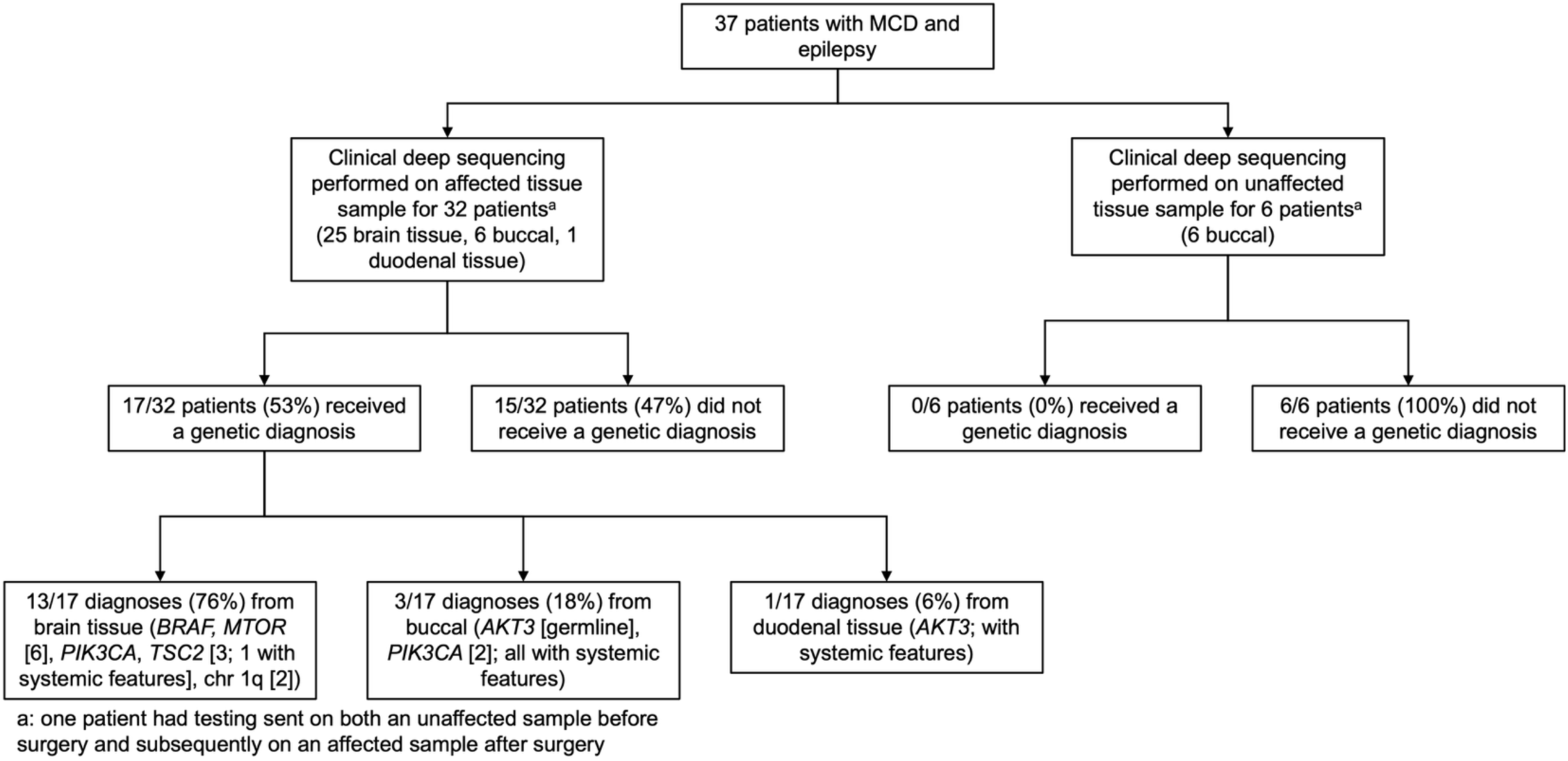
Study Workflow.

For the 32 individuals with clinical deep sequencing performed on an affected tissue sample, testing was performed on brain tissue for 25 individuals (1 with systemic features), buccal for 6 individuals (all with systemic features), and duodenal tissue for 1 individual (with systemic features). Most of these individuals had one clinical deep sequencing test designed for individuals with suspected non-oncologic disorders of mosaicism: 14 had the CME panel, 8 had the SO panel, 2 had a custom panel combining the CME and SO panel genes (of which 1 also had a prior SO panel on an unaffected tissue sample, see below), and 1 had a custom CME/Rasopathies panel. An additional 2 individuals had the BRIGHTseq Beacon test designed for individuals with known or suspected tumors. The remaining 5 individuals had multiple clinical deep sequencing tests performed on affected samples: 2 had the SO panel and Oncopanel (of which 1 also had a chromosomal microarray (CMA) performed on an affected sample), 1 had the CME panel and Oncopanel, 1 had a custom CME/SO panel and BRIGHTseq Beacon, and 1 had the SO panel and subsequently the CME panel.

The diagnostic yield for clinical deep sequencing performed on an affected tissue sample was 53% (17/32). Of the 17 individuals who received genetic diagnoses, 13/17 (76%) had testing performed on brain tissue (1 with systemic features; **Table 2**) and 4/17 (24%) had testing performed on non-brain tissue (3 from buccal and 1 from duodenal tissue, all with systemic features; **Table 3**). Clinical deep sequencing identified P/LP mosaic variants in *AKT3* (1 individual, duodenal tissue [systemic features]), *BRAF* (1 individual, brain tissue), *MTOR* (6 individuals, brain tissue), *PIK3CA* (1 individual, brain tissue; 2 individuals, buccal [systemic features]), *TSC2* (3 individuals, brain tissue [1 with systemic features)], as well as mosaic chr 1q gain (2 individuals, brain tissue). The VAF of the diagnostic mosaic variants was reported by the clinical laboratory for 9 individuals and ranged from 1- 17%. For 6 individuals with diagnostic mosaic variants, sequencing was also performed on a comparator blood sample. The mosaic variant was not detected in the corresponding blood sample for 5 individuals (via Sanger sequencing for 3 and deep sequencing for 2) and was detected in the corresponding blood sample for 1 individual (via Sanger sequencing; *PIK3CA* variant initially identified using a buccal sample in an individual with systemic features). A pathogenic germline variant was also identified in *AKT3* (1 individual, buccal [systemic features]).

**Table 2:**
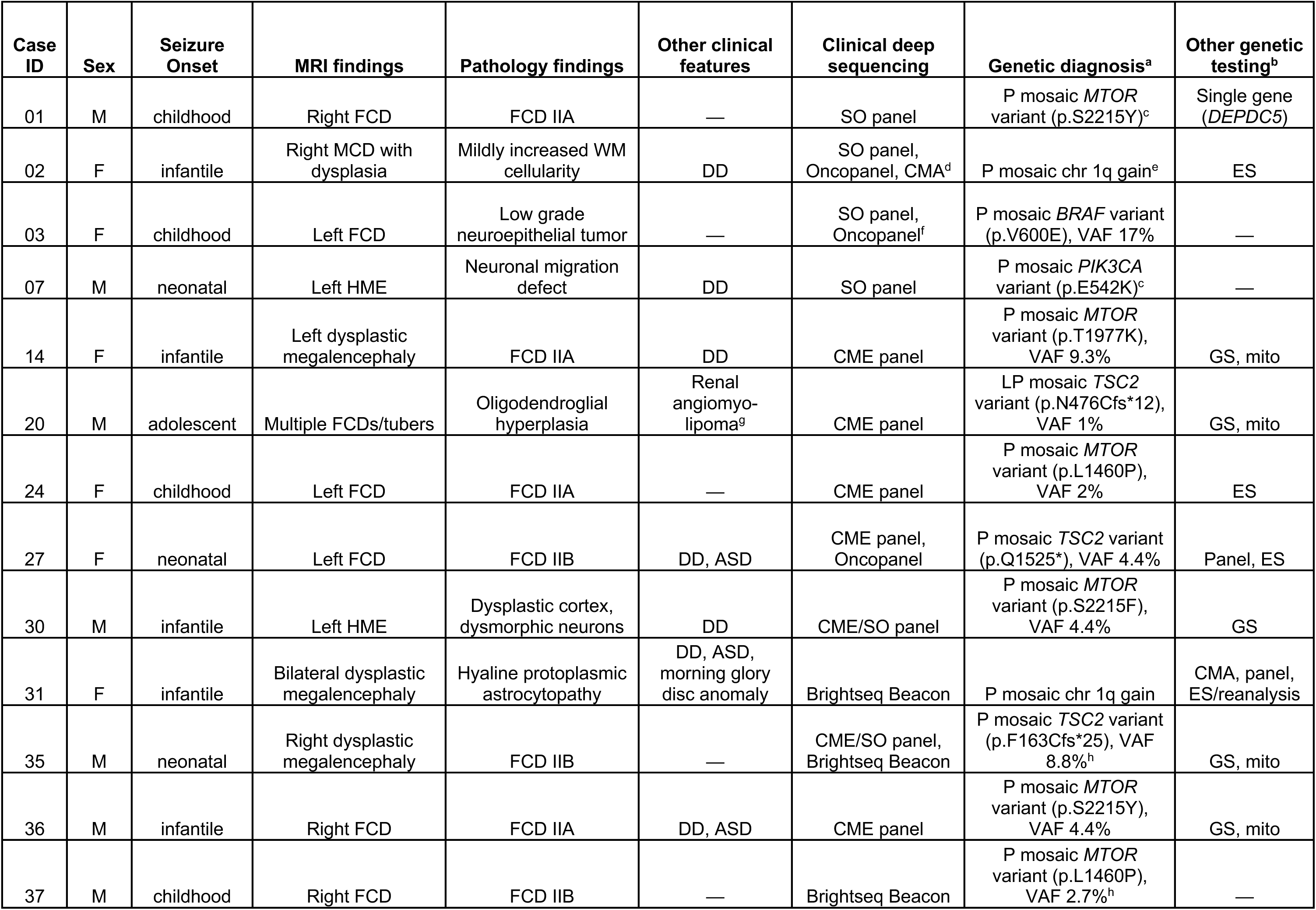
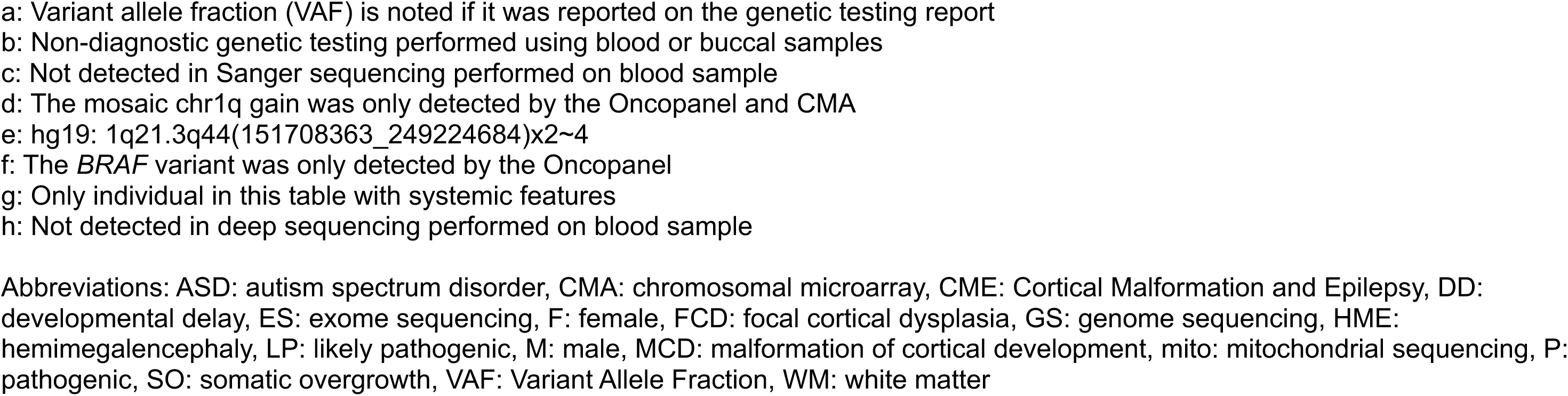
Summary of Diagnostic Findings Detected in Brain Tissue Samples.

| Case ID | Sex | Seizure Onset | MRI findings | Pathology findings | Other clinical features | Clinical deep sequencing | Genetic diagnosis<sup>a</sup> | Other genetic testing<sup>b</sup> |
| --- | --- | --- | --- | --- | --- | --- | --- | --- |
| 01 | M | childhood | Right FCD | FCD IIA | — | SO panel | P mosaic *MTOR* variant (p.S2215Y)<sup>c</sup> | Single gene (*DEPDC5*) |
| 02 | F | infantile | Right MCD with dysplasia | Mildly increased WM cellularity | DD | SO panel, Oncopanel, CMA<sup>d</sup> | P mosaic chr 1q gain<sup>e</sup> | ES |
| 03 | F | childhood | Left FCD | Low grade neuroepithelial tumor | — | SO panel, Oncopanel<sup>f</sup> | P mosaic *BRAF* variant (p.V600E), VAF 17% | — |
| 07 | M | neonatal | Left HME | Neuronal migration defect | DD | SO panel | P mosaic *PIK3CA* variant (p.E542K)<sup>c</sup> | — |
| 14 | F | infantile | Left dysplastic megalencephaly | FCD IIA | DD | CME panel | P mosaic *MTOR* variant (p.T1977K), VAF 9.3% | GS, mito |
| 20 | M | adolescent | Multiple FCDs/tubers | Oligodendroglial hyperplasia | Renal angiomyolipoma<sup>g</sup> | CME panel | LP mosaic *TSC2* variant (p.N476Cfs*12), VAF 1% | GS, mito |
| 24 | F | childhood | Left FCD | FCD IIA | — | CME panel | P mosaic *MTOR* variant (p.L1460P), VAF 2% | ES |
| 27 | F | neonatal | Left FCD | FCD IIB | DD, ASD | CME panel, Oncopanel | P mosaic *TSC2* variant (p.Q1525*), VAF 4.4% | Panel, ES |
| 30 | M | infantile | Left HME | Dysplastic cortex, dysmorphic neurons | DD | CME/SO panel | P mosaic *MTOR* variant (p.S2215F), VAF 4.4% | GS |
| 31 | F | infantile | Bilateral dysplastic megalencephaly | Hyaline protoplasmic astrocytopathy | DD, ASD, morning glory disc anomaly | Brightseq Beacon | P mosaic chr 1q gain | CMA, panel, ES/reanalysis |
| 35 | M | neonatal | Right dysplastic megalencephaly | FCD IIB | — | CME/SO panel, Brightseq Beacon | P mosaic *TSC2* variant (p.F163Cfs*25), VAF 8.8%<sup>h</sup> | GS, mito |
| 36 | M | infantile | Right FCD | FCD IIA | DD, ASD | CME panel | P mosaic *MTOR* variant (p.S2215Y), VAF 4.4% | GS, mito |
| 37 | M | childhood | Right FCD | FCD IIB | — | Brightseq Beacon | P mosaic *MTOR* variant (p.L1460P), VAF 2.7%<sup>h</sup> | — |

- a: Variant allele fraction (VAF) is noted if it was reported on the genetic testing report - b: Non-diagnostic genetic testing performed using blood or buccal samples - c: Not detected in Sanger sequencing performed on blood sample - d: The mosaic chr1q gain was only detected by the Oncopanel and CMA - e: hg19: 1q21.3q44(151708363\_249224684)x2~4 - f: The *BRAF* variant was only detected by the Oncopanel - g: Only individual in this table with systemic features - h: Not detected in deep sequencing performed on blood sample
Abbreviations: ASD: autism spectrum disorder, CMA: chromosomal microarray, CME: Cortical Malformation and Epilepsy, DD: developmental delay, ES: exome sequencing, F: female, FCD: focal cortical dysplasia, GS: genome sequencing, HME: hemimegalencephaly, LP: likely pathogenic, M: male, MCD: malformation of cortical development, mito: mitochondrial sequencing, P: pathogenic, SO: somatic overgrowth, VAF: Variant Allele Fraction, WM: white matter

**Table 3:** Summary of Diagnostic Findings Detected in Non-Brain Tissue Samples.

| Case ID | Sex | Seizure Onset | MRI findings | Pathology findings | Systemic features | Other clinical features | Clinical deep sequencing | Sample used | Genetic diagnosis<sup>a</sup> | Other genetic testing<sup>b</sup> |
| --- | --- | --- | --- | --- | --- | --- | --- | --- | --- | --- |
| 04 | M | N/A | Bilateral PMG/PVNH | — | MCAP-like syndrome | DD | SO panel | Buccal | P germline *AKT3* variant (p.R465W) | Karyotype |
| 06 | M | childhood | Bilateral PMG | — | MCAP syndrome | DD, ASD | SO panel | Buccal | LP mosaic *PIK3CA* variant (p.E418K)<sup>c</sup> | CMA |
| 13 | F | neonatal | Right HME | FCD IIA | MCAP syndrome | DD | SO panel | Duodenal tissue | P mosaic *AKT3* variant (p.E17K) | — |
| 22 | M | infantile | Bilateral dysplastic megalencephaly | — | CLOVES syndrome | DD | SO panel | Buccal | P mosaic *PIK3CA* variant (p.E110del)<sup>d</sup> | Single gene (*PIK3CA*), ES |

a: Variant allele fraction (VAF) is listed if it was reported on the genetic testing report
b: Non-diagnostic genetic testing performed using blood or buccal samples
c: Detected in Sanger sequencing performed on a blood sample
d: Not detected in Sanger sequencing performed on a blood sample
Abbreviations: ASD: autism spectrum disorder, CLOVES: Congenital Lipomatous Overgrowth, Vascular Malformations, Epidermal Nevis, Spinal/Skeletal Anomalies/Scoliosis, DD: developmental delay, F: female, FCD: focal cortical dysplasia, HME: hemimegalencephaly, M: male, MCAP: Megalencephaly-Capillary Malformation, P: pathogenic, PMG: polymicrogyria, PVNH: periventricular nodular heterotopia, SO: somatic overgrowth

Four individuals had multiple clinical deep sequencing tests performed on affected samples and received genetic diagnoses. For 2 of those individuals, the diagnostic variants were detected by both clinical deep sequencing tests: a mosaic *TSC* variant was detected by both a CME panel and Oncopanel and another mosaic *TSC2* variant was detected by both a custom CME/SO panel and BRIGHTseq Beacon. For the other 2 individuals, the diagnostic variants were not detected by one of the clinical deep sequencing tests: a mosaic *BRAF* variant was detected by Oncopanel but not by a SO panel (*BRAF* is not included on the SO panel) and a mosaic 1q gain was detected by Oncopanel (and a CMA) but not by a SO panel (CNVs are not detectable by the SO panel).

Apart from one individual with an FCD suspected on MRI who was ultimately found to have a low grade neuroepithelial tumor on pathology and for whom clinical deep sequencing identified a mosaic variant in *BRAF*, the individuals who received genetic diagnoses all had diagnostic variants involving the mTOR pathway (**Figure 2**). Moreover, apart from two individuals with MRI findings of bilateral PMG in the setting of MCAP/MCAP-like syndrome, the individuals who received genetic diagnoses all had MRI findings representing a spectrum of dysplasias (**Figure 3**).

**Figure 2:**
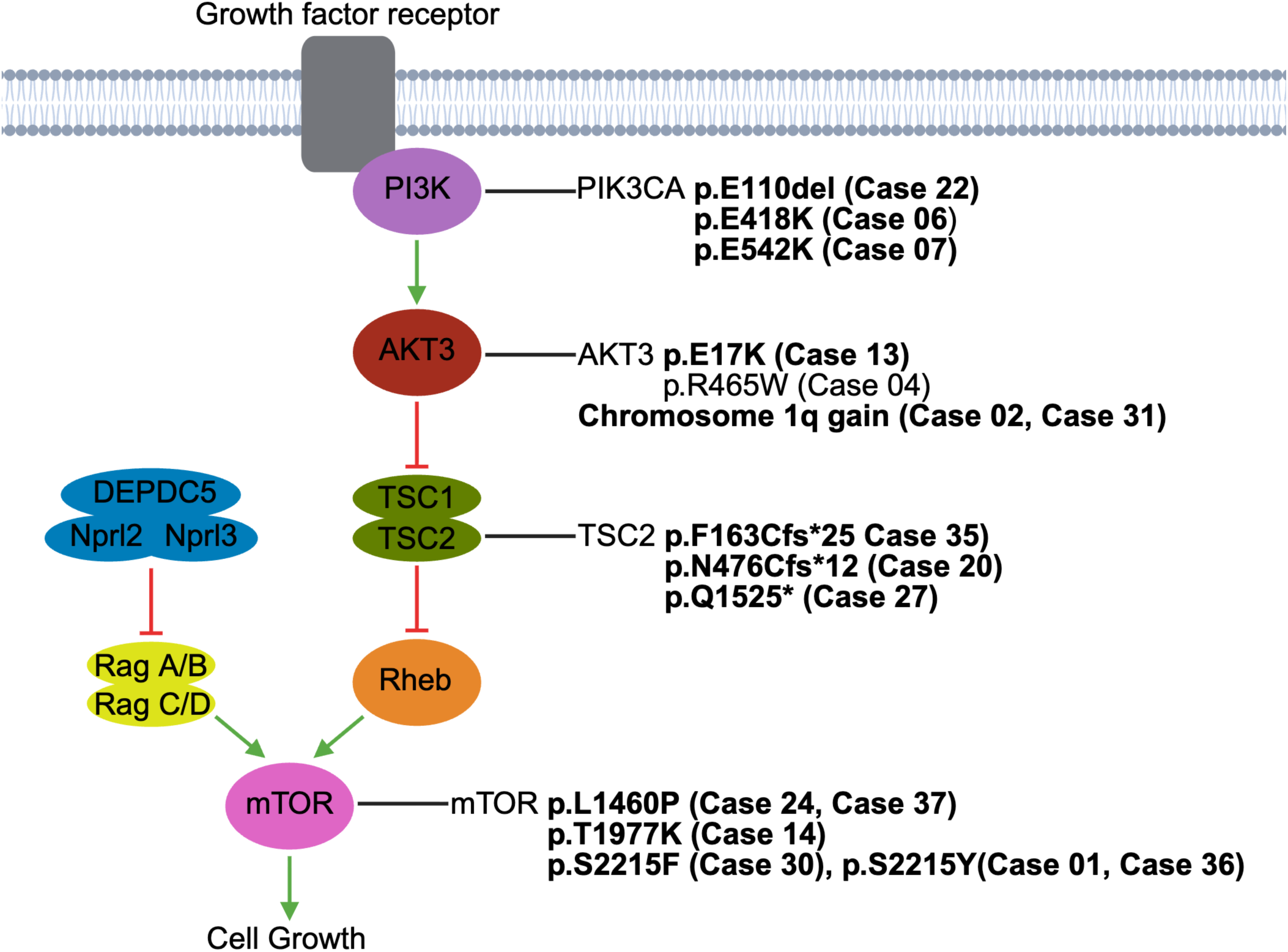
mTOR Pathway and Pathogenic Variants. Schematic of the mTOR pathway annotated with the Pathogenic/Likely Pathogenic variants identified in the cohort. The only variant not annotated is the *BRAF* mosaic variant in Case 03. Mosaic variants are shown in boldface. Created using BioRender.

**Figure 3:**
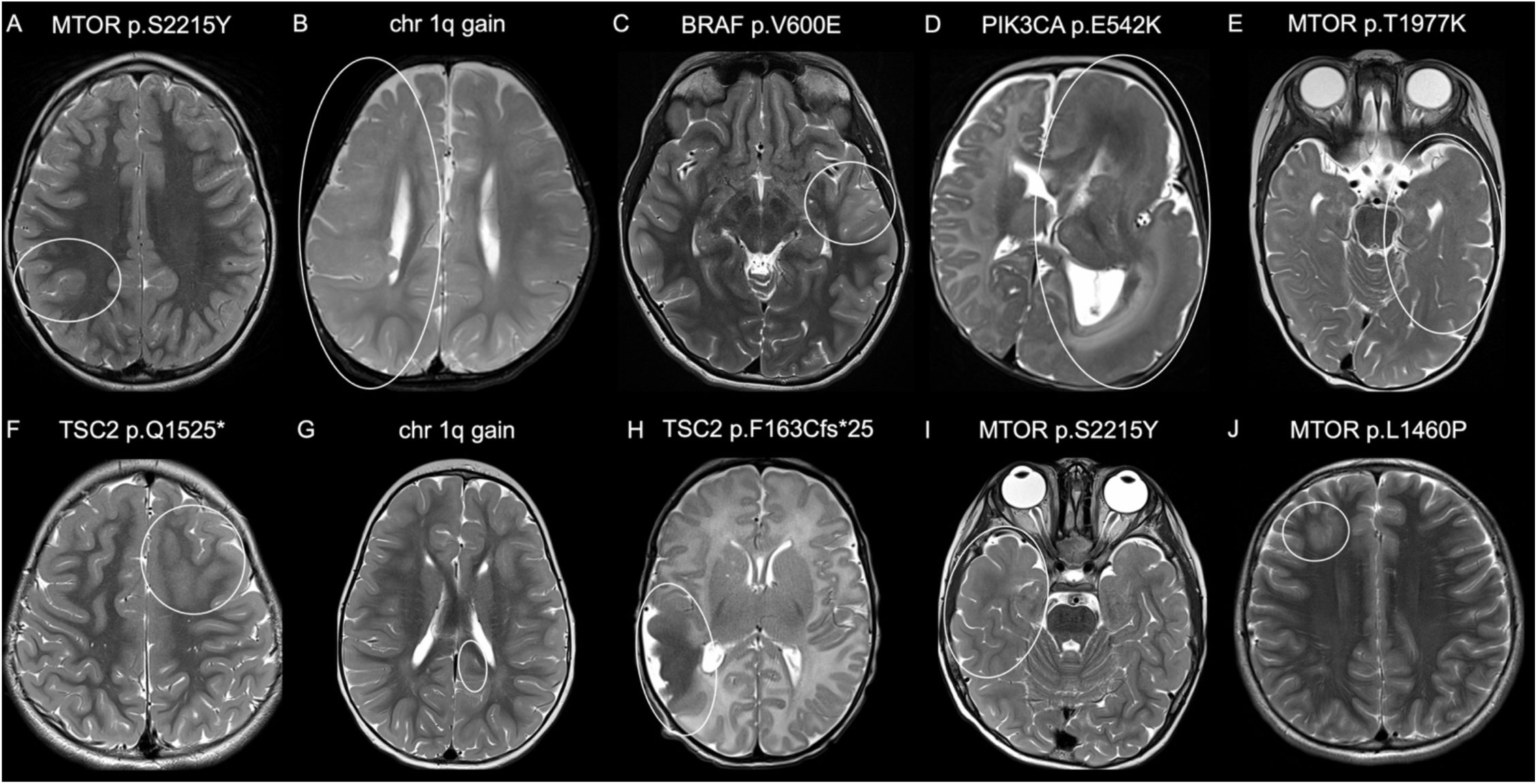
Brain Imaging Findings in Individuals with Pathogenic Mosaic Variants. Axial T2 weighted images from Cases 01 (A), 02 (B), 03 (C), 07 (D), 14 (E), 27 (F), 31 (G), 35 (H), 36 (I), and 37 (J). Circles denote areas of MCD. The identified mosaic variant for each case is noted.

For the 6 individuals with clinical deep sequencing performed on an unaffected tissue sample, testing was performed on buccal samples (none with systemic features). The individuals all had one clinical deep sequencing test designed for individuals with suspected non-oncologic disorders of mosaicism: 5 had the SO panel (of which 1 also had a subsequent custom CME/SO panel on an affected sample, see above) and 1 had the CME panel. The diagnostic yield for clinical deep sequencing performed on an unaffected sample was 0% (0/6). Details for the individuals who did not receive a genetic diagnosis from clinical deep sequencing tests are available in **Supplementary Table 1**.

Overall, the diagnostic yield was higher when clinical deep sequencing was performed using an affected tissue sample vs an unaffected tissue sample (17/32 (53%) vs 0/6 (0%); p=0.016). In addition, the diagnostic yield was associated with pathology findings (p=0.017). Notably, the diagnostic yield was 89% (8/9) for individuals with pathology findings of FCD II; these individuals all had clinical deep sequencing performed on an affected tissue sample.

Most individuals (29/37 (78%)) also had other genetic testing focused on germline variant detection (individuals could have had more than one other genetic test). These other genetic tests were performed on blood or buccal samples and were non-diagnostic (**Table 2**, **Table 3, Supplementary Table 1**). Chromosome-level testing was performed for 11 individuals, including 10 who had a CMA and 1 who had a karyotype. Focused gene-level testing was performed for 12 individuals, including 9 who had one or more multi-gene panels and 6 who had single gene/single condition testing. Genomic sequencing was performed for 24 individuals, including 18 who had exome sequencing, 13 who had genome sequencing, and 13 who had mitochondrial sequencing.

### Clinical Utility

All of the genetic diagnoses made by clinical deep sequencing had clinical utility for the individuals and their families (**Table 4**). Notably, all had potential precision therapy implications, including mTOR pathway and *BRAF* inhibitors. The genetic diagnoses led to new subspecialist referrals for 6/17 individuals (35%) and to additional workup (laboratory tests or imaging) for 6/17 individuals (35%). For all cases, the genetic diagnoses informed prognosis beyond the prognosis associated with the clinical features, largely based on the type of diagnostic variant (mosaic vs germline) and the existing literature on outcomes of reported individuals with the same or similar variants. In addition, for all cases the genetic diagnoses informed reproductive counseling for the parents of the individual as well as for the individuals themselves. As one example, Case 036 is a male child with infantile-onset DRE in the setting of FCD on MRI, confirmed as FCD IIa on pathology after surgical resection, as well as comorbid developmental delay and autism. He had non-diagnostic genome and mitochondrial sequencing performed on unaffected samples and was found to have a pathogenic mosaic *MTOR* variant at a VAF of 4.4% on a CME panel performed on an affected brain tissue sample. The genetic diagnosis informed potential precision therapies (mTOR pathway inhibitors), informed prognosis based on the variant type (mosaic and distinct from germline Smith-Kingsmore syndrome) and on reported individuals with the same or similar variant, and informed recurrence risk (negligible risk for parents for future pregnancies and low risk for the individual given likely brain-limited).

**Table 4:** Clinical Utility of Diagnostic Findings.

| **Case ID** | **Gene/**<br>**Genomic**<br>**Region** | **Potential**<br>**precision**<br>**therapy** | **New**<br>**referral** | **New labs/**<br>**imaging** | **Inform**<br>**prognosis** | **Inform**<br>**recurrence**<br>**risk** |
| --- | --- | --- | --- | --- | --- | --- |
| 01 | *MTOR* | Yes | — | — | Yes | Yes |
| 02 | chr 1q | Yes | — | — | Yes | Yes |
| 03 | *BRAF* | Yes | Yes | Yes | Yes | Yes |
| 04 | *AKT3*<sup>a</sup> | Yes | Yes | Yes | Yes | Yes |
| 06 | *PIK3CA* | Yes | Yes | Yes | Yes | Yes |
| 07 | *PIK3CA* | Yes | — | — | Yes | Yes |
| 13 | *AKT3* | Yes | — | — | Yes | Yes |
| 14 | *MTOR* | Yes | — | — | Yes | Yes |
| 20 | *TSC2* | Yes | Yes | Yes | Yes | Yes |
| 22 | *PIK3CA* | Yes | — | — | Yes | Yes |
| 24 | *MTOR* | Yes | — | — | Yes | Yes |
| 27 | *TSC2* | Yes | — | Yes | Yes | Yes |
| 30 | *MTOR* | Yes | — | — | Yes | Yes |
| 31 | chr 1q | Yes | Yes | — | Yes | Yes |
| 35 | *TSC2* | Yes | Yes | Yes | Yes | Yes |
| 36 | *MTOR* | Yes | — | — | Yes | Yes |
| 37 | *MTOR* | Yes | — | — | Yes | Yes |

a: germline variant

## DISCUSSION

In this study, we report our use of clinical deep sequencing to diagnose pathogenic mosaic variants in individuals with MCDs, primarily cortical dysplasia, and epilepsy. Overall, we find high diagnostic yield (>50%) and clinical utility of clinical deep sequencing for this population when testing is able to be performed on an affected tissue sample.

The importance of performing testing on an affected tissue sample, and how we defined an affected tissue sample, reflects our understanding of the time and place the somatic mutation event leading to the relevant phenotype occurs during development. If a mosaic variant arises prior to gastrulation, it may be present across the three germ layers and thus detectable across multiple tissues. In some MCD phenotypes without systemic (non-CNS) features, such as double cortex syndrome, deep sequencing has detected mosaic variants in blood or saliva samples, supporting a relatively early somatic mutation event.^2, 33^ Similar findings have been reported for mosaic variants detected in individuals with MCD phenotypes with systemic features, including various overgrowth syndromes like MCAP.^34^ In other MCD phenotypes without systemic features, including the dysplasia phenotypes in our cohort, deep sequencing of collectively hundreds of individuals in the research setting has detected mosaic variants in brain tissue samples but usually not in available non-brain tissue samples like blood, suggesting that the mosaic variants arose after gastrulation.^19, 23, 35, 36^ If a mosaic variant arises after gastrulation but before neurogenesis, it may be present and detectable across ectodermal tissues including buccal samples. However, prior studies and our current findings demonstrate that mosaic variants in isolated dysplasia phenotypes are usually not detected in non-brain ectodermal tissue like buccal samples, suggesting that the mosaic variants arose during neurogenesis.^37^ If a mosaic variant arises during neurogenesis, it may be brain-limited and thus detectable only in brain tissue. While exceptions have been reported, most studies in isolated dysplasia phenotypes support the somatic mutation event occurring in a neural progenitor, with examples reported of cases with the mosaic variant detected in neuronal but not non-neuronal cells.^36, 38^ Thus, in our cohort focused on individuals with dysplasia phenotypes, we conservatively defined an affected tissue sample as a brain tissue sample for individuals with a MCD phenotype without systemic features and as a brain tissue sample or an involved non-brain tissue sample (based on the reported systemic features) for individuals with a MCD phenotype with systemic features.

Our diagnostic yield was over 50% when clinical deep sequencing was performed on an affected tissue sample compared to 0% on an unaffected tissue sample. Currently, relatively few clinical laboratories offer accredited deep sequencing tests to detect mosaic variants in non-oncologic disorders of mosaicism.^19, 39^ In a prior heterogenous cohort of individuals referred to the Clinical Genomics Laboratory at Washington University School of Medicine in St. Louis (the same laboratory used in this study) for suspected disorders of non-oncologic mosaicism, similarly high diagnostic yields were reported when testing was performed on an affected tissue sample vs low diagnostic yields when testing was performed on an unaffected tissue sample.^37, 40^ Of note, these prior studies considered buccal samples as affected if the individual had disease phenotypes in the head and reported high diagnostic yields when clinical deep sequencing was performed on buccal samples for individuals with disease phenotypes in the head, including brain malformations. Although limited clinical information from clinical requisitions were available for the individuals included in these prior studies, for individuals reported to have isolated dysplasia phenotypes, diagnostic variants were only detected when clinical deep sequencing was performed on affected fresh (frozen) or FFPE tissue samples (4/7 cases; all mosaic and not present in corresponding blood sample) and not detected when clinical deep sequencing was performed on blood or buccal samples (0/7 cases), consistent with our findings.^37^ Prior studies from a hospital in the Czech Republic reported diagnostic yields of 18% (5/28; all mosaic and not present in corresponding blood sample) and 29% (10/35; 7/10 mosaic) when clinical deep sequencing (panel targeting 43 genes) was performed on affected brain tissue samples from individuals with MCDs, also largely dysplasia phenotypes.^41, 42^ Thus, clinical deep sequencing has high diagnostic yield for individuals with MCDs, especially dysplasia phenotypes, when performed on an appropriate affected sample. We highlight the importance of rigorous phenotyping for individuals with MCDs, incorporating as available imaging, pathology, and a detailed physical exam, to classify the type of MCD and the presence or absence of systemic features. When selecting and performing clinical genetic testing, such rigorous phenotyping has important implications for the predicted molecular genetic landscape of the underlying disorder and for which samples should be considered affected.

In our cohort, the affected tissue samples were primarily brain tissue samples from individuals with MCDs without systemic features who underwent resective surgery for DRE. Thus, for individuals with isolated MCDs, an affected tissue sample was only available for individuals who underwent resective surgery and genetic diagnosis was only possible post-surgery. Proof-of-concept exists in the research setting for innovative methods to detect mosaic variants earlier in the epilepsy journey using less invasive sources of brain-derived DNA, including the small amounts of genomic DNA from brain cells adherent to stereo-EEG electrodes and cell free DNA from CSF.^14, 19, 43–46^ While these methods require optimization compared to the gold standard of genomic DNA from resected brain tissue, they hold promise for future diagnostic use in individuals with MCDs and epilepsy who do not undergo surgical resection and/or prior to surgical resection.

Implementation of clinical deep sequencing for individuals with MCDs and epilepsy in our institution required multidisciplinary engagement and coordination between epilepsy, neurosurgery, neuropathology, genetic counseling, laboratory medicine, and the genetic testing laboratory. For most individuals, clinical deep sequencing was covered by payors as part of routine clinical care. We report high clinical utility of returning results of deep sequencing to individuals, their families, and treating clinicians, which to the best of our knowledge has not been assessed in this population. Even though the genetic diagnoses were made post-surgery for most individuals, the diagnoses impacted multiple aspects of clinical utility for the individuals and their families.^19^ All of the genetic diagnoses had implications for potential precision therapies, and establishing a genetic diagnosis allows eligibility for clinical trials or compassionate use of existing or emerging precision therapies like mTOR pathway inhibitors.^47^ As with all genetic testing, pre-test consent/genetic counseling and post-test genetic counseling is important to ensure families understand the implications of genetic testing and the genetic results, including recurrence risk implications and interpretation of Variants of Uncertain Significance (VUS). For a mosaic variant, the risk to parents of the affected individual is negligible for future pregnancies as the mosaic variant is presumed to have occurred post-zygotically in the affected individual, although rare exceptions have been reported of more complex mechanisms (e.g., post-zygotic rescue of mitotic errors).^48^ However, the recurrence risk to the affected individual depends on the tissue distribution of the identified mosaic variant and whether the variant is truly somatic vs gonosomal (i.e., present in a subset of somatic and germ cells). For individuals in our cohort without systemic features for whom a mosaic variant was detected in brain tissue, we counseled that the individual is unlikely to carry the variant in cells outside of the brain including gametes, while acknowledging that without testing these cells we cannot absolutely know. For individuals in our cohort with systemic features for whom a mosaic variant was detected in brain or non-brain tissue, we counseled that the individual may carry the variant in gametes and thus have a non-negligible risk of passing the variant to future children.

Limitations of our study include the single-center nature with a relatively small number of individuals. Given that deep sequencing for this population has largely been performed in the research setting, we believe it is important to report on our implementation of such testing in the clinical setting in a cohort of individuals with detailed clinical information, including imaging and pathology findings. Most but not all of the individuals in our cohort had clinical germline genetic testing, which can inform surgical workup especially in pediatric populations.^42, 49^ It is possible that the few individuals in our study who did not have germline genetic testing and did not receive a genetic diagnosis from clinical deep sequencing carry a diagnostic germline variant. The limited demographic diversity of our cohort reflects the demographics of our hospital but not necessarily that of individuals with MCD and epilepsy more broadly. Future studies should investigate the diagnostic yield of clinical deep sequencing and relevant associations with diagnostic yield in larger and more diverse cohorts of individuals with epilepsy and MCD. In addition, our analysis of clinical utility is based on clinician-perceived utility abstracted from the EMR. To scale implementation of clinical deep sequencing into routine clinical care for individuals with MCD and epilepsy, and for individuals with non-oncologic disorders of somatic mosaicism more broadly, future studies should investigate facilitators and barriers to implementation, particularly given the limited numbers of clinical genetics providers, as well as investigate the clinician and individual/parent-perceived utility of such testing. Finally, ongoing development of clinically accredited tests for mosaic variants and efforts to establish consensus criteria for classification of mosaic variants in non-oncologic disorders of mosaicism, which require modification from criteria for classification of germline variants and thus far have been published for mosaic variants in *AKT3*, *MTOR*, *PIK3CA*, and *PIK3R2*, will further advance implementation.^23, 50^

In conclusion, clinical deep sequencing, when performed using an affected tissue sample, has high diagnostic utility for individuals with MCDs, especially dysplasia phenotypes, and epilepsy. Our findings support implementation of clinical deep sequencing for this population, especially as the genetic diagnoses have implications for emerging precision therapies, including mTOR pathway inhibitors, and clinical trials.

## Conflict of Interest Disclosures

The other authors report no conflicts of interest.

## Funding/Support

AMD was supported by the Boston Children’s Hospital Translational Research Program, a Boston Children’s Hospital Office of Faculty Development/Basic & Clinical Translational Research Executive Committees Faculty Career Development Fellowship, and by NIH/NINDS K23 NS140397. EY and AP were supported by NIH/NINDS U24 NS137481. This research was conducted with support from the BCH Children’s Rare Disease Collaborative and International Precision Child Health Partnership as well as the BCH Rosamund Stone Zander and Hansjoerg Wyss Translational Neuroscience Center Repository Core for Neurological Disorders, which is also supported by the IDDRC (NIH P50HD105351). This work was supported by the National Institutes of Health under award numbers K23 NS140397, U24 NS137481, and P50 HD105351. This manuscript is the result of funding in whole or in part by the National Institutes of Health (NIH). It is subject to the NIH Public Access Policy. Through acceptance of this federal funding, NIH has been given a right to make this manuscript publicly available in PubMed Central upon the Official Date of Publication, as defined by NIH.

## Additional Acknowledgments

We thank the individuals and their families who made this study possible. We also thank the clinicians who referred and returned results of clinical deep sequencing tests as part of clinical care.

## Notes

### Competing Interest Statement

The authors have declared no competing interest.

### Author Declarations

The Boston Children's Hospital (BCH) Institutional Review Board (IRB) approved this retrospective chart review study with a waiver of informed consent. A subset of the individuals included in this study, including those whose imaging is shown in Figure 3, are enrolled in BCH IRB-approved research studies for which the individual or their parent provided written informed consent. These include the Rosamund Stone Zander and Hansjoerg Wyss Translational Neuroscience Center Repository Core for Neurological Disorders, the Children's Rare Disease Collaborative Unexplained Epilepsies study, and the Gene-STEPS study.

