## Supplementary for "Clinical deep sequencing to diagnose pathogenic mosaic variants in malformations of cortical development and epilepsy"

**Supplementary Table 1: Summary of Non-Diagnostic Findings**

| Case ID | Sex | Seizure Onset | MRI findings | Pathology findings | Systemic features | Other clinical features | Clinical deep sequencing | Sample used | Affected sample used | Other genetic testing[a] |
|---|---|---|---|---|---|---|---|---|---|---|
| 05 | F | infantile | Left MCD with FCD | FCD IIA | — | DD, ADHD | CME panel | Brain tissue | Yes | Panel, ES |
| 08 | M | infantile | Left parietal FCD/tuber | N/A | — | DD, ADHD, learning disability, sensory processing difficulties | SO panel | Buccal | No | CMA, single gene (*TSC1/2*), panels |
| 09 | F | infantile | Right frontal FCD | N/A | — | DD, ADHD | SO panel | Buccal | No | Panel, ES, GS, mito |
| 10 | F | childhood | Right HME | N/A | Dysmorphic features, heterochromic iridis, dermatologic and size differences between right and left body | DD, ID | SO panel | Buccal | Yes | CMA, Fragile X, Waarden-burg syndrome |
| 11 | M | N/A | Right HME | N/A | PROS | DD, learning disability | SO panel | Buccal | Yes | CMA, ES |
| 12 | F | infantile | Left frontal MCD including FCD and PVNH | Mild malformative features, focal disorganization, oligodendroglial hyperplasia; PVNH; | — | — | CME panel | Brain tissue | Yes | — |
| 15 | F | infantile | Suspected right temporal FCD | Oligodendroglial hyperplasia; Hippocampal sclerosis, dentate gyrus bilamination and dispersion | — | Learning disabilities | CME panel | Brain tissue | Yes | GS |

| | | | | | | | | | | |
|---|---|---|---|---|---|---|---|---|---|---|
| 16 | F | childhood | Suspected left frontotemporal FCD and right MTS | Possible mild MCD | — | ASD, ADHD, anxiety, Type I Diabetes | CME panel | Brain tissue | Yes | CMA, ES, GS, mito |
| 17 | M | childhood | Left HME with PMG | N/A | — | DD | SO panel | Buccal | No | — |
| 18 | F | neonatal | Bilateral cortical dysplasia | N/A | Left hemibody hemiplegia, left leg cutis marmorata telangiectasia congenital | DD, regression, strabismus and esotropia | SO panel, CME panel | Buccal | Yes | CMA, single gene (*IKBKG*), panel, ES |
| 19 | M | infantile | Left cortical dysplasia | MCD | — | DD | CME panel | Brain tissue | Yes | GS, mito |
| 21 | F | neonatal | Left HME | Focal microcolumnar arrangement, rare dysmorphic neurons | — | DD, ASD | SO panel | Buccal | No | — |
| 23 | M | childhood | Right temporal FCD and MTS | Oligodendroglial hyperplasia, HS | — | — | CME/Rasopathies panel | Brain tissue | Yes | Panel, GS, mito |
| 25 | F | adolescent | Left temporal FCD | FCD IA | — | ASD, ID, ADHD | CME panel | Brain tissue | Yes | ES, mito |
| 26 | M | childhood | MSGP | N/A | — | DD, mixed sleep apnea | CME panel | Buccal | No | CMA, ES |
| 28 | M | childhood | Left frontal FCD | Oligodendroglial hyperplasia | — | DD, regression | CME panel | Brain tissue | Yes | CMA, panel, ES, mito |
| 29 | M | infantile | Left > Right MCD including left frontal FCD and PMG | Possible MCD | — | DD | CME panel | Brain tissue | Yes | CMA, panels, ES, GS, mito, Angleman/ PWS |

| | | | | | | | | | | |
|---|---|---|---|---|---|---|---|---|---|---|
| 32 | F | infantile | Left HME | Mild MCD/FCD IA | — | Duplex kidney | SO panel (buccal), CME/SO panel (brain) | Buccal and brain tissue | Initially No, Subsequently Yes | GS, mito |
| 33 | F | infantile | Suspected left temporal FCD | FCD IC | — | DD, regression | CME panel | Brain tissue | Yes | ES, GS, mito |
| 34 | F | infantile | Suspected left frontal FCD | MCD | — | Prematurity (33 weeks), DD | CME Panel | Brain tissue | Yes | — |

a: Non-diagnostic genetic testing performed using blood or buccal samples

Abbreviations: ADHD: attention deficit hyperactivity disorder, ASD: autism spectrum disorder, CMA: chromosomal microarray, CME: Cortical Malformation and Epilepsy, DD: developmental delay, ES: exome sequencing, F: female, FCD: focal cortical dysplasia, GS: genome sequencing, HME: hemimegalencephaly, ID: intellectual disability, M: male, MCD: malformation of cortical development, mito: mitochondrial sequencing, MSGP: microcephaly with simplified gyral pattern, MTS: mesial temporal sclerosis, PMG: polymicrogyria, PROS: PIK3CA-related overgrowth spectrum, PVNH: periventricular nodular heterotopia, SO: somatic overgrowth

**Supplementary Figure 1: Additional Brain Imaging Findings in Individuals with Pathogenic Mosaic Variants.**

Additional axial images from Cases 01 (A), 02 (B), 03 (C), 07 (D), 14 (E), 27 (F), 31 (G), 35 (H), 36 (I), and 37 (J). Circles denote areas of MCD. (A) and (G) are axial FLAIR images and the remainder are axial T1 MPRAGE images.

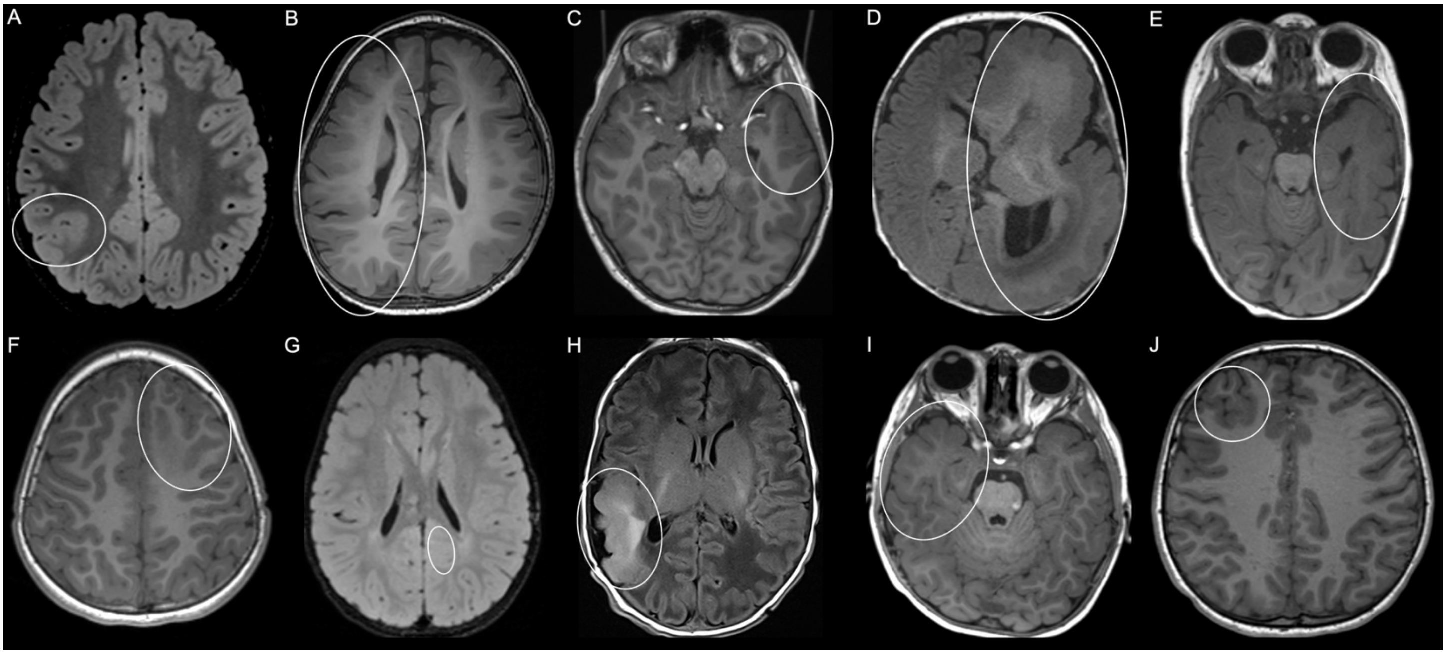
